# Effectiveness of Monovalent Rotavirus Vaccine Among Young Children in Pakistan: a Test-Negative Case-Control Evaluation

**DOI:** 10.1101/2025.05.10.25327376

**Authors:** Syed Asad Ali, Shazia Sultana, Atif Riaz, Mohammad Tahir Yousafzai, Aneeta Hotwani, Furqan Kabir, Muhammad Nasir Rana, Yasin Alvi, Saadia Khan, Khalid Iqbal, Mubashir Ahmed, Muhammad Haroon Hamid, Amir Muhammad, Inayat Ullah Afridi, Arit Parkash, Nasir Saleem Saddal, Jamal Raza, Muhammad Hayat Bozdar, Zafar Iqbal Channa, Zareef Uddin Khan, Muhammad Naeem Rajput, Sohail Raza Shaikh, Asad Ali, Mathew Esona, Rashi Gautam, Umesh D. Parashar, Jacqueline E. Tate, Margaret M. Cortese, Muhammad Ahmed Kazi

## Abstract

**Background:** Pakistan ranks among top 4 countries with greatest number of rotavirus deaths among young children. To reduce the burden of severe rotavirus disease, Pakistan began rolling out the monovalent oral rotavirus vaccine, Rotarix (RV1), in late 2017 and reached all areas by April 2018, using a 2-dose schedule at ages 6 and 10 weeks.

**Methods:** During April 2018 through March 2023, we performed active surveillance for acute rotavirus gastroenteritis resulting in intravenous hydration at 9 hospital in 3 provinces among children eligible to have received rotavirus vaccine. Cases were children whose stool sample tested positive for rotavirus by enzyme immunoassay and controls were those testing negative. Unconditional logistic regression was used to estimate the effectiveness of 2 doses of RV1 vs 0 doses in reducing likelihood of hospital care for rotavirus disease.

**Results:** Over 2900 children were enrolled and had an analyzable vaccination record obtained. Based on 517 rotavirus cases and 1310 controls, the effectiveness of 2 RV1 doses among children aged 22 weeks–11 months was 33% (95% CI 12, 49); the effectiveness was 45% (95% CI 21, 62) among those better nourished by weight-for-age z-score (WAZ>-2). Among children aged 1 year, the effectiveness was 24% (95% CI -20, 52). A wide array of genotypes were detected and nearly 40% of cases had >1 G and/or P genotype identified. Vaccine effectiveness point estimates were lowest for those with genotype P[4] detected vs those with P[8] or P[6] detected.

**Conclusion:** 2 RV1 doses appear to provide fair protection in this very high burden setting. Additional vaccination strategies should continue to be pursued for children in this and similar locations.

## INTRODUCTION

The implementation of rotavirus vaccine in national immunization programs has been demonstrated to be life-saving, and to substantially reduce the morbidity from rotavirus disease among young children across regions as assessed, for example, by the reduction in the proportion of hospitalizations for acute gastroenteritis diarrhea that are due to rotavirus [1,2]. Before rotavirus vaccine introduction, Pakistan ranked within the top 4 countries with the highest burden of rotavirus-associated deaths among children aged <5 years, with an estimated 14,700 rotavirus deaths annually and rotavirus mortality rate of 67.6 per 100,000 children [3].

Four different oral rotavirus vaccines have demonstrated efficacy in clinical trials and have been pre-qualified by WHO [4]. It is valuable, therefore, to better understand how well different rotavirus vaccines perform under routine use, especially in locations where the severe rotavirus disease burden is very high. These highest burden settings are also the ones where the challenges for performance of an oral rotavirus vaccine administered in infancy are greatest, with factors such as concomitant administration with oral polio vaccine, high prevalence of malnutrition and environmental enteric enteropathy, and high levels of transplacental antibodies among the factors believed to hinder vaccine-induced protection [5].

In January 2017, Pakistan began the roll-out of the monovalent human rotavirus vaccine, Rotarix (RV1, GSK, Rixensart Belgium) for its children with all areas providing vaccine by March/April 2018. The first and second doses are recommended to be given at ages 6 and 10 weeks, along with other routine immunizations including oral polio vaccine. The objective of our evaluation was to estimate the effectiveness of the RV1 vaccine series in reducing the likelihood of receiving hospital care for moderate or severe acute rotavirus gastroenteritis, using a test-negative case control design. This information is expected to help guide public health policy and vaccination strategies.

## METHODS

### Enrollment

Active surveillance for rotavirus gastroenteritis in children aged <5 years had been previously established at selected hospitals in Pakistan [6]. After vaccine introduction, surveillance activities were expanded to additional hospitals at different time points and included procedures to be able to measure vaccine effectiveness (VE) [7] through a joint venture that included the Expanded Program of Immunization, the Ministry of National Health Services, Regulations and Coordination, and partner hospitals. The VE evaluation network ultimately included 9 hospitals in 3 provinces (Sindh, Punjab and Khyber Paktunkwa) (Figure). Monday through Saturday, generally 9am to 2pm, dedicated study staff screened children cared for in the emergency departments, pediatric medical wards, and/or diarrhea treatment units to identify and enroll eligible children. Children eligible for enrollment were those aged ≥ 4 months and eligible by date of birth to have received ≥1 dose of RV1 before illness (depending on vaccine introduction date in the area [Map/Table]), presenting with diarrhea (≥3 looser-than-normal stools in a 24-hour period during the illness; onset ≤7 days before presentation) as a main reason for care, and receipt of intravenous hydration. Children were ineligible for enrollment if they had bloody diarrhea, had already been enrolled for the same illness episode or had already been hospitalized for >48 hours before diarrhea onset. Written informed consent was obtained from parents/legal guardian willing to have their child participate. Because of the COVID-19 pandemic, enrollment was suspended during approximately mid-March 2020 through late May 2020.

A standardized data collection form was used to collect information from parent/guardians on the child’s illness, family/household characteristics (e.g., number of children in the household, type of housing material, source of drinking water (Supplemental Table 1), and vaccination information (see below). A whole stool sample was to be collected as soon as possible after presentation and stored at 4°C. Information on the child’s course of illness (i.e. maximum temperature during the first 24 hours of hospital care, date of discharge) was also recorded. At the time of discharge (or as close to this time as possible), the child’s height and weight were to be measured by trained study staff.

### Vaccination Information

Parents were queried as to whether child had ever received any vaccines, were asked the name and location of clinics that had provided any vaccines, and were asked to provide the parent-held copy of any vaccination record so that a photograph could be made. For parents who did not have the record with them during hospital care, follow-up efforts were made with the parents to retrieve and photograph the record. For children whose parents reported the child had received vaccinations after the birth doses but who did not have a record or for whom the record was unclear, attempts were made whenever feasible to visit the vaccination clinic and review any manual vaccination register to obtain the vaccination information. Parents who did not have a vaccine record for their child but who reported that their child had not received any vaccines subsequent to any birth doses were considered to have had no RV1 doses. For children enrolled in the last months of the evaluation from Sindh province who did not have a parent-held record to photograph, staff attempted to obtain the child’s vaccination information using a mobile phone application that had become available in the area. Dates of vaccine doses as indicated in the photographs/register/app were recorded on the data form; project coordinators also reviewed the photographs and forms for accuracy of vaccination information and adjudicated records when necessary. All staff were blinded to stool EIA-result when vaccine records were reviewed.

### Laboratory procedures

Stool samples were tested for rotavirus antigen by enzyme immunoassay (EIA) at the Infectious Diseases Research Laboratory (IDRL) at Aga Khan University, except that samples collected at Mayo Hospital were tested by their own hospital laboratory. Samples from Karachi sites were sent to IDRL on daily basis and samples from outside Karachi were shipped regularly through courier service; cold chain was maintained. IDRL and Mayo Hospital tested samples using the ProSpecT Rotavirus Test (Oxoid), following the manufacturer’s instructions. After EIA testing at IDRL, any remaining sample was stored at -20°C.

At IDRL, rotavirus EIA-positive samples were extracted using Qiagen Viral RNA kit (following manufacturer’s instructions) and analyzed for strain characterization using a multiplex one-step reverse transcriptase PCR (RT-PCR) methodology to ascertain the G- and P-types of the strains [8]. Samples with >1 G and/or P-type based on results from multiplex assay testing were subsequently tested with single-plex RT-PCR for genotype confirmation [8]. EIA-positive samples that did not have G and P type identifiable were classified as non-typeable strains. A subset of samples with G1P[8] detected were additionally tested to determine if the strain was RV1 strain using real-time RT-PCR and RV1-strain specific primers [9,10]. Nearly all EIA-positive samples from Mayo Hospital were not available to be genotyped at IDRL.

### Statistical analyses

Age at admission and at receipt of each RV1 dose was calculated. For VE analyses, an RV1 dose was counted if it had been received ≥14 days prior to presentation at hospital. The VE of 2 doses of RV1 vs 0 doses among children receiving hospital care for acute gastroenteritis was calculated as (1 – odds ratio) x 100%. The odds ratios for receipt of 2 RV1 doses among rotavirus cases compared with test-negative controls was determined using unconditional logistic regression; models a priori included month of birth, year of birth, calendar quarter of hospital care, year of hospital care, and hospital; age category (2-month intervals) was also included for children aged <1 year. We calculated the VE for children aged ≥ 22 weeks to minimize any residual confounding by age; cases and controls aged <22 weeks were not included in the analyses. Case patients and controls eligible to be included in VE estimates were also compared through bivariate analysis on household/family characteristics (Supplemental Table 1); those factors with p-values <.05 were assessed for possible confounding in each model by backward elimination and retained if the VE point estimate changed by ≥ 2.5 percentage points. Weight-for-age category (see below) was also assessed for possible confounding in the models. Analyses planned a priori included VE by age (e.g., age 22 weeks–11 months, age 12 months–23 months, age 22 weeks–23 months, using separate models), by age and case illness severity (using separate models, with case illness severity categorized by a modified scoring system [Supplemental Table 2] and length of hospital stay), by age and case genotype (separate models; cases and controls from Mayo Hospital were excluded because genotype was unavailable for nearly all cases) and by age and nutritional status. For the latter, each child was classified based on the 2006 WHO Child Growth Standards indices [11]: weight-for-age (WA), height/length-for-age (HA) and weight-for-height (WFH) z-scores whereby scores <-2 indicates at least moderate malnutrition. We obtained VE estimates by nutritional status (z-scores <-2 vs ≥ -2) by using an interaction term in the models. When models were based on smaller numbers of cases, (i.e∼60-70), Firth correction was incorporated [12]. Post-hoc, we examined VE by other available clinical severity information (e.g., reported level of dehydration) and additional nutritional analysis (e.g., both WA and HA z-score ≥-2). In sensitivity analyses, we excluded the 2.6% of children aged <2 years with WA considered out-of-range by WHO from the VE by WAZ analysis [13]. Similarly, for the VE by HAZ analysis, we excluded the 3.8% of children with HA considered out-of-range. All analyses were conducted using Stata software vs. 13 (StataCorp, TX).

For the VE estimates of 2 vs 0 doses, children who had received only 1 RV1 dose or who received 1 or 2 doses but the exact number was unknown were excluded. To avoid misclassification of rotavirus vaccine status, we excluded from analysis children whose vaccination record indicated they had received DTP or OPV doses before admission but the card format did not have a labelled row for the vaccinator to record doses of rotavirus vaccine administered (i.e., card formats had not yet been updated to include labelled space for rotavirus vaccine) and rotavirus vaccine doses had not otherwise been recorded on the card. When possible, before excluding, a clinic visit was made to determine if the clinic had manual logs whereby a child’s rotavirus vaccine status could be confirmed. A post-hoc VE analysis was performed including a subset of these children excluded from the main analysis (Supplemental Material).

The project was approved by the Ethical Review Committees of all the participating hospitals and the National Bioethics Committee of Pakistan. It was deemed to be public health evaluation by the Human Subjects Committee at CDC.

## RESULTS

During April 2018 through March 2023, 6454 children were eligible for enrollment and a stool sample was collected and tested by EIA in 4833 (75%). Among those children with an EIA result, rotavirus vaccination information was available on 78% (3774/4833) overall [83% of infants and 76% of children aged 12-23 months], including 83% of the total 1037 children whose stool tested rotavirus-positive and 77% of the 3796 whose stool tested rotavirus-negative.

### Children with an analyzable vaccination record

Overall, 38% of children with an EIA result and an analyzable vaccination record were from Sindh province, 53% from Punjab and 9% from Khyber Pakhtunkhwa. Stool was collected ≤1 day from presentation for 91% of them. Among the 97% of children with a known outcome and who were discharged home; 32% were discharged on the same calendar day as arrival, 24% were discharged the day after admission, and 44% had a longer stay. Among children aged <12 months, the WA z-score was <-2 in 40% of those who tested rotavirus-positive and in 51% of those who tested negative (Supplemental Table 3). Among those aged 12-23 months, the proportions were 30% and 44%, respectively.

The median (25^th^, 75^th^ percentile) age at receipt of RV1 dose 1 was 8.0 weeks (6.8, 10.7) and at dose 2 was 13.5 weeks (11.7, 17.0). Among 1485 children aged 22 weeks−11 months who tested rotavirus-negative and for whom vaccination information was available, 1066 (72%) had received 2 RV1 doses, 163 (11%) had received 1 dose, 12 (1%) had received 1 or 2 doses but exact number was unknown, and 244 (16%) had received no doses. Among the 589 who tested rotavirus-positive, the values were 386 (66%), 70 (12%), 2 (<1%), and 131 (22%), respectively. Of the children who had received 2 RV1 doses, 99.8% received OPV doses on the same dates.

### Vaccine effectiveness overall and by nutritional status

Among children aged 22 weeks−11 months, the overall VE for 2 RV1 doses was 33% (95% CI 13, 49) and for age 12−23 months was 24% (95% CI -20, 52) (Table1). With both groups combined, the VE for children aged 22 week – 23 months was 31% (95% CI 13, 45). Stratified by WAZ score, the VE point estimate was higher among better nourished infants, 45% (95% CI 21, 62) as compared to malnourished infants 23% (95% CI -13, 48) (Table 2, Supplemental Table 4). By HAZ score, the VE results were similar among infants who were not stunted and those stunted. Using a separate model post-hoc, the VE among infants whose WAZ and HAZ score were both ≥-2 was 55% (95% CI 27, 72) (Supplemental Table 4). Among infants, the VE of 1 RV1 dose vs 0 doses was 22% (95% CI -13, 47).

**Table 1.**
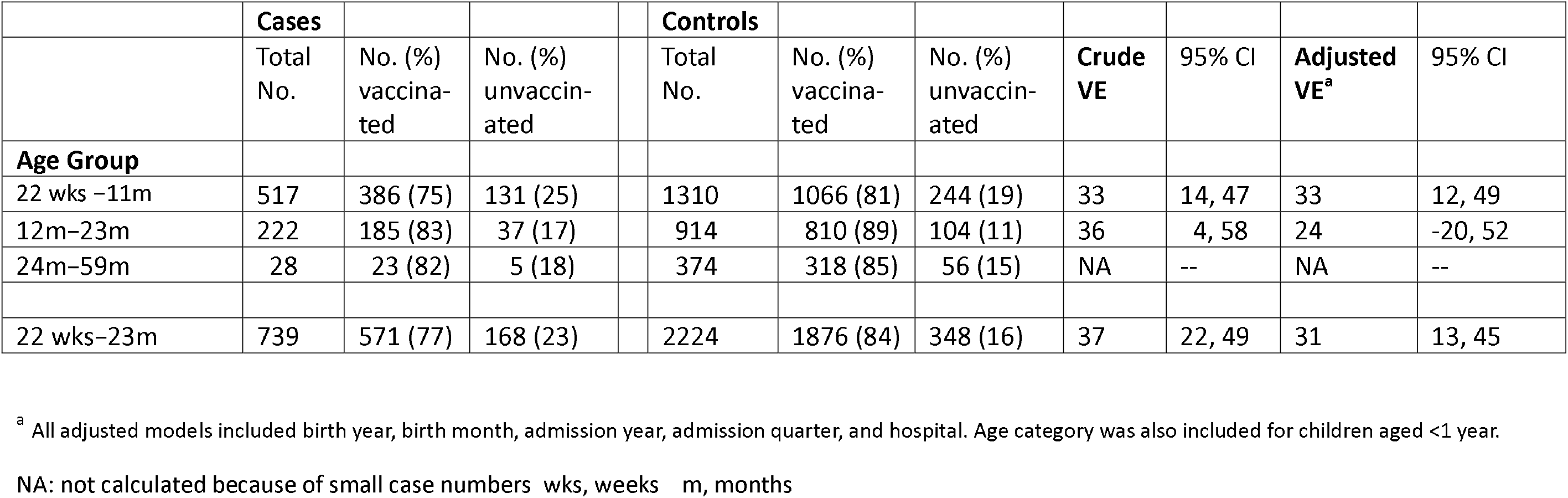
Vaccine effectiveness of 2 doses of RV1 vs 0 doses, by age group - Pakistan, 4/2018 – 3/2023.

|  | **Cases** |  |  |  | **Controls** |  |  |  |  |  |  |
| --- | --- | --- | --- | --- | --- | --- | --- | --- | --- | --- | --- |
|  | Total<br>No. | No. (%)<br>vaccina-<br>ted | No. (%)<br>unvaccin-<br>ated |  | Total<br>No. | No. (%)<br>vaccina-<br>ted | No. (%)<br>unvaccin-<br>ated | **Crude**<br>**VE** | 95% CI | **Adjusted**<br>**VE<sup>a</sup>** | 95% CI |
| **Age Group** |  |  |  |  |  |  |  |  |  |  |  |
| 22 wks –11m | 517 | 386 (75) | 131 (25) |  | 1310 | 1066 (81) | 244 (19) | 33 | 14, 47 | 33 | 12, 49 |
| 12m–23m | 222 | 185 (83) | 37 (17) |  | 914 | 810 (89) | 104 (11) | 36 | 4, 58 | 24 | -20, 52 |
| 24m–59m | 28 | 23 (82) | 5 (18) |  | 374 | 318 (85) | 56 (15) | NA | -- | NA | -- |
| 22 wks–23m | 739 | 571 (77) | 168 (23) |  | 2224 | 1876 (84) | 348 (16) | 37 | 22, 49 | 31 | 13, 45 |

<sup>a</sup> All adjusted models included birth year, birth month, admission year, admission quarter, and hospital. Age category was also included for children aged <1 year.
NA: not calculated because of small case numbers wks, weeks m, months

**Table 2.** Vaccine effectiveness of 2 doses of RV1 vs 0 doses, by age group and nutritional status - Pakistan, 4/2018 – 3/2023.

|  | Cases |  |  | Controls |  |  |  |  |  |  |  |
| --- | --- | --- | --- | --- | --- | --- | --- | --- | --- | --- | --- |
|  | Total No.<br>(% of total<br>cases) | No. (%)<br>vaccinated | No. (%)<br>unvaccinated |  | Total<br>No. (%<br>of total<br>controls<br>) | No. (%)<br>vaccinated | No. (%)<br>unvaccinated |  | VE<sup>a</sup> | 95% CI | p-value<br>interaction<br>term |
| Age 22 weeks<br>–11m |  |  |  |  |  |  |  |  |  |  |  |
| WA z-score ≥-2 | 313 (61) | 236 (75) | 77 (25) |  | 633 (48) | 537 (85) | 96 (15) |  | 45 | 21, 62 | 0.21 |
| WA z-score <-2 | 204 (39) | 150 (74) | 54 (26) |  | 676 (52) | 528 (78) | 148 (22) |  | 23 | -13, 48 |  |
| HA z-score ≥-2 | 305 (59) | 240 (79) | 65 (21) |  | 711 (54) | 605 (85) | 106 (15) |  | 34 | 4, 54 | 0.89 |
| HA z-score <-2 | 211 (41) | 145 (69) | 66 (31) |  | 598 (46) | 460 (77) | 138 (23) |  | 36 | 7, 56 |  |
| WH z-score ≥-2 | 348 (67) | 258 (74) | 90 (26) |  | 785 (60) | 643 (82) | 142 (18) |  | 38 | 13, 55 | 0.55 |
| WH z-score <-2 | 168 (33) | 127 (76) | 41 (24) |  | 521 (40) | 420 (81) | 101 (19) |  | 26 | -15, 53 |  |
| Age 12m –23m |  |  |  |  |  |  |  |  |  |  |  |
| WA z-score ≥-2 | 154 (69) | 132 (86) | 22 (14) |  | 508 (56) | 452 (89) | 56 (11) |  | 5 | -73, 48 | 0.27 |
| WA z-score <-2 | 68 (31) | 53 (78) | 15 (22) |  | 405 (44) | 357 (88) | 48 (12) |  | 43 | -15, 72 |  |
| HA z-score ≥-2 | 117 (53) | 102 (87) | 15 (13) |  | 515 (56) | 463 (90) | 52 (10) |  | -4 | -105, 48 | 0.28 |
| HA z-score <-2 | 105 (47) | 83 (79) | 22 (21) |  | 398 (44) | 346 (87) | 52 (13) |  | 38 | -17, 67 |  |
| WH z-score ≥-2 | 176 (79) | 147 (84) | 29 (16) |  | 561 (61) | 498 (89) | 63 (11) |  | 21 | -36, 55 | 0.92 |
| WH z-score <-2 | 46 (21) | 38 (83) | 8 (17) |  | 352 (39) | 311 (88) | 41 (12) |  | 25 | -85, 69 |  |

<sup>a</sup>All models included birth year, birth month, admission year, admission quarter, and hospital. Age category was also included for children aged <1 year.

Among children aged 12−23 months at illness, all VE estimates by nutritional category included zero and for two assessments (WAZ and HAZ), there was wide discrepancy in the point estimates with low values in the better nourished children and high values in the malnourished (Table 2, Supplemental Table 5). Results from the sensitivity analyses for VE by nutritional status were similar to the original results (Supplemental Tables 4 and 5).

Nearly all (85%) of infants in the analysis had a modified Vesikari severity score ≥11 so VE results for cases in this category were similar as the overall VE (Table 3). VE point estimates appeared modestly higher in a few higher severity subset categories (i.e., score ≥ 13, and cases classified as lethargic accounting for only 13% of cases). Similarly, among children aged 12-23 months, point estimates were generally low (and CI included zero) except for score ≥15 where VE was 42% (95% CI -4, 66), accounting for 38% of cases (Table 3).

**Table 3.**
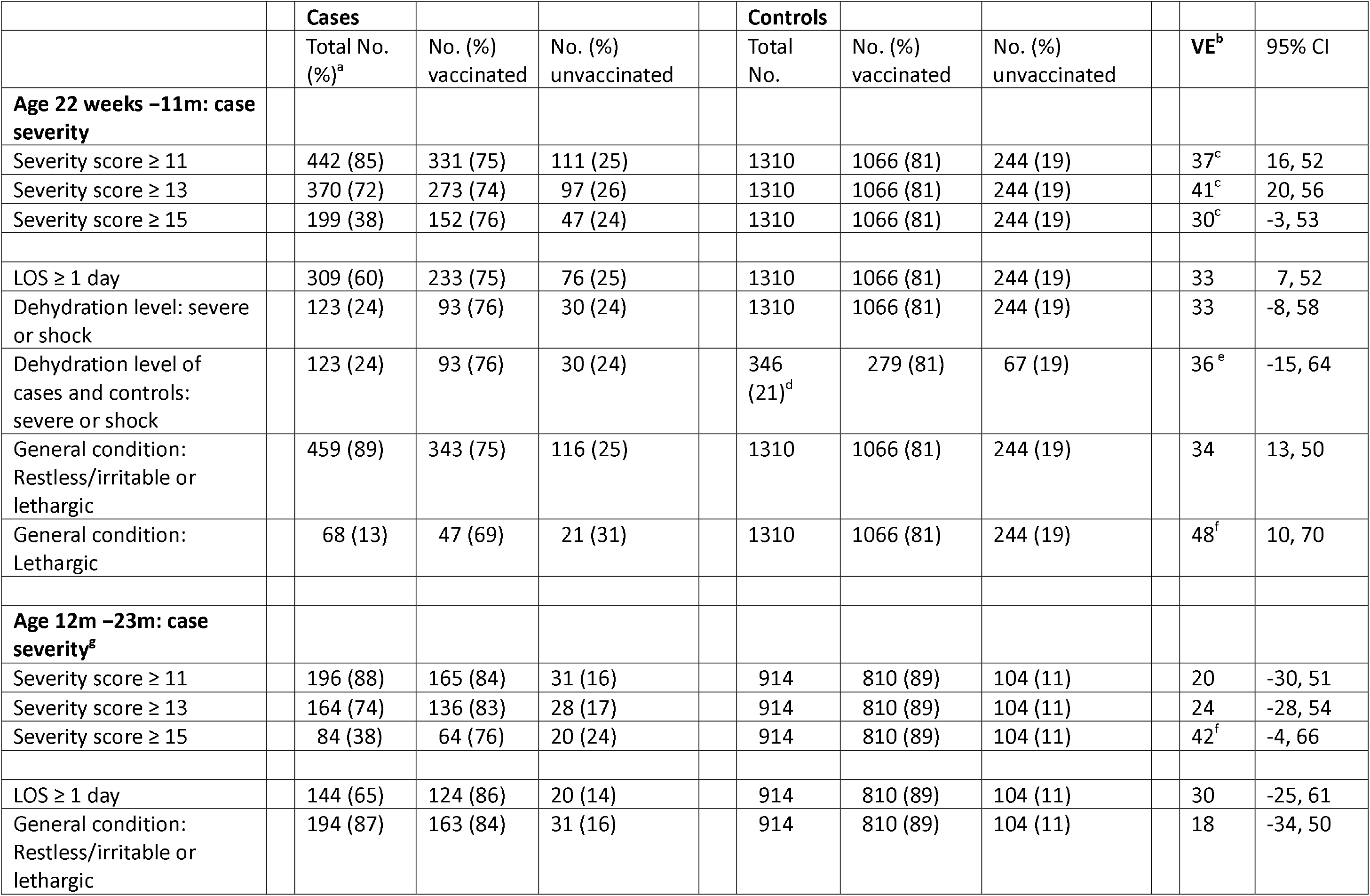
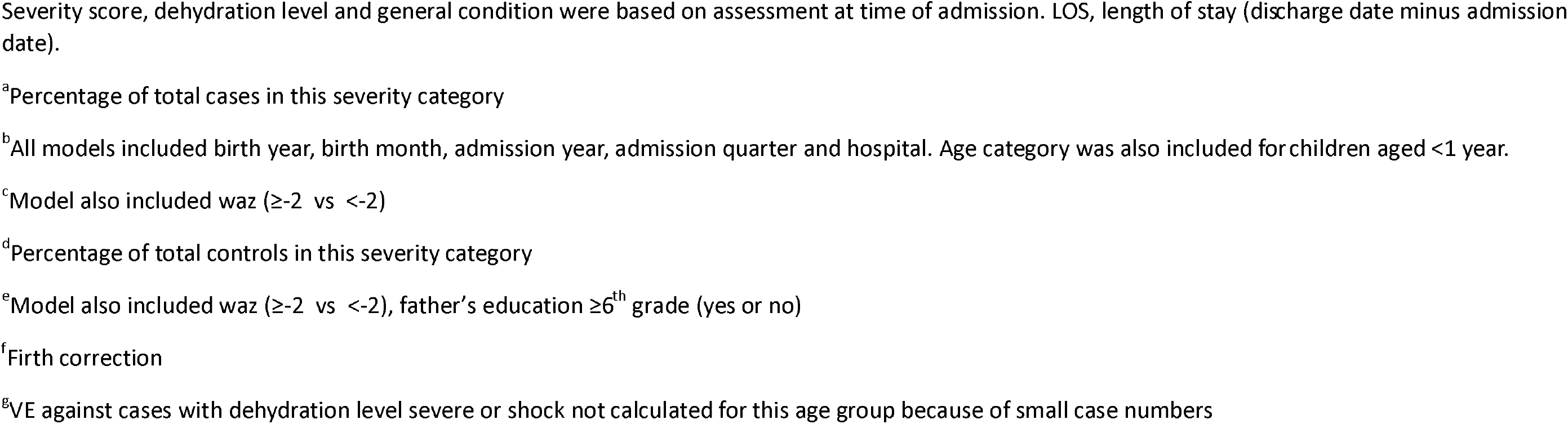
Vaccine effectiveness of 2 doses of RV1 vs 0 doses, by age group and case severity status-Pakistan, 4/2018 – 3/2023.

|  | **Cases** |  |  |  | **Controls** |  |  |  |  |
| --- | --- | --- | --- | --- | --- | --- | --- | --- | --- |
|  | Total No.<br>(%)<sup>a</sup> | No. (%)<br>vaccinated | No. (%)<br>unvaccinated |  | Total<br>No. | No. (%)<br>vaccinated | No. (%)<br>unvaccinated | **VE<sup>b</sup>** | 95% CI |
| **Age 22 weeks –11m: case severity** |  |  |  |  |  |  |  |  |  |
| Severity score ≥ 11 | 442 (85) | 331 (75) | 111 (25) |  | 1310 | 1066 (81) | 244 (19) | 37<sup>c</sup> | 16, 52 |
| Severity score ≥ 13 | 370 (72) | 273 (74) | 97 (26) |  | 1310 | 1066 (81) | 244 (19) | 41<sup>c</sup> | 20, 56 |
| Severity score ≥ 15 | 199 (38) | 152 (76) | 47 (24) |  | 1310 | 1066 (81) | 244 (19) | 30<sup>c</sup> | -3, 53 |
| LOS ≥ 1 day | 309 (60) | 233 (75) | 76 (25) |  | 1310 | 1066 (81) | 244 (19) | 33 | 7, 52 |
| Dehydration level: severe or shock | 123 (24) | 93 (76) | 30 (24) |  | 1310 | 1066 (81) | 244 (19) | 33 | -8, 58 |
| Dehydration level of cases and controls: severe or shock | 123 (24) | 93 (76) | 30 (24) |  | 346 (21)<sup>d</sup> | 279 (81) | 67 (19) | 36<sup>e</sup> | -15, 64 |
| General condition: Restless/irritable or lethargic | 459 (89) | 343 (75) | 116 (25) |  | 1310 | 1066 (81) | 244 (19) | 34 | 13, 50 |
| General condition: Lethargic | 68 (13) | 47 (69) | 21 (31) |  | 1310 | 1066 (81) | 244 (19) | 48<sup>f</sup> | 10, 70 |
| **Age 12m –23m: case severity<sup>g</sup>** |  |  |  |  |  |  |  |  |  |
| Severity score ≥ 11 | 196 (88) | 165 (84) | 31 (16) |  | 914 | 810 (89) | 104 (11) | 20 | -30, 51 |
| Severity score ≥ 13 | 164 (74) | 136 (83) | 28 (17) |  | 914 | 810 (89) | 104 (11) | 24 | -28, 54 |
| Severity score ≥ 15 | 84 (38) | 64 (76) | 20 (24) |  | 914 | 810 (89) | 104 (11) | 42<sup>f</sup> | -4, 66 |
| LOS ≥ 1 day | 144 (65) | 124 (86) | 20 (14) |  | 914 | 810 (89) | 104 (11) | 30 | -25, 61 |
| General condition: Restless/irritable or lethargic | 194 (87) | 163 (84) | 31 (16) |  | 914 | 810 (89) | 104 (11) | 18 | -34, 50 |

Severity score, dehydration level and general condition were based on assessment at time of admission. LOS, length of stay (discharge date minus admission date).
<sup>a</sup>Percentage of total cases in this severity category
<sup>b</sup>All models included birth year, birth month, admission year, admission quarter and hospital. Age category was also included for children aged <1 year.
<sup>c</sup>Model also included waz ( $\geq -2$ vs $< -2$ )
<sup>d</sup>Percentage of total controls in this severity category
<sup>e</sup>Model also included waz ( $\geq -2$ vs $< -2$ ), father's education $\geq 6^{\text{th}}$ grade (yes or no)
<sup>f</sup>Firth correction
<sup>g</sup>VE against cases with dehydration level severe or shock not calculated for this age group because of small case numbers

### Genotype distribution and vaccine effectiveness by genotype

Among children aged 22 weeks–11 months in the VE analysis for 2 doses vs 0 doses, genotyping was performed on for 485/491 (99%) rotavirus cases enrolled from hospitals other than Mayo Hospital. Of the 485 samples with genotype results, rotavirus was classified as G and P non-typeable in 6 (1%). Five different G types (G1, G2, G3, G9, G12), 3 different P types (P[4,] P[6], P[8]), and 13 different single G and P genotype combinations were identified (Figure 2, Supplemental Table 6). In 38% (180/479 cases with at least G or P-typeable), >1 G and/or P type was detected (in 25%, only >1 G type was identified, in 9% only >1 P type was identified, and in 4% >1 G and P type were identified). The most common G and P genotype combinations were G9P[4], G12P[6], and G1P[8]. More than half (268/485 [56%]) contained neither G1 nor P[8]. Of the 161 cases aged <24 months where G1P[8] was detected and for whom vaccination status was available, testing for RV1 strain was performed on 56 (35%) samples and none were positive.

**Figure 1:**
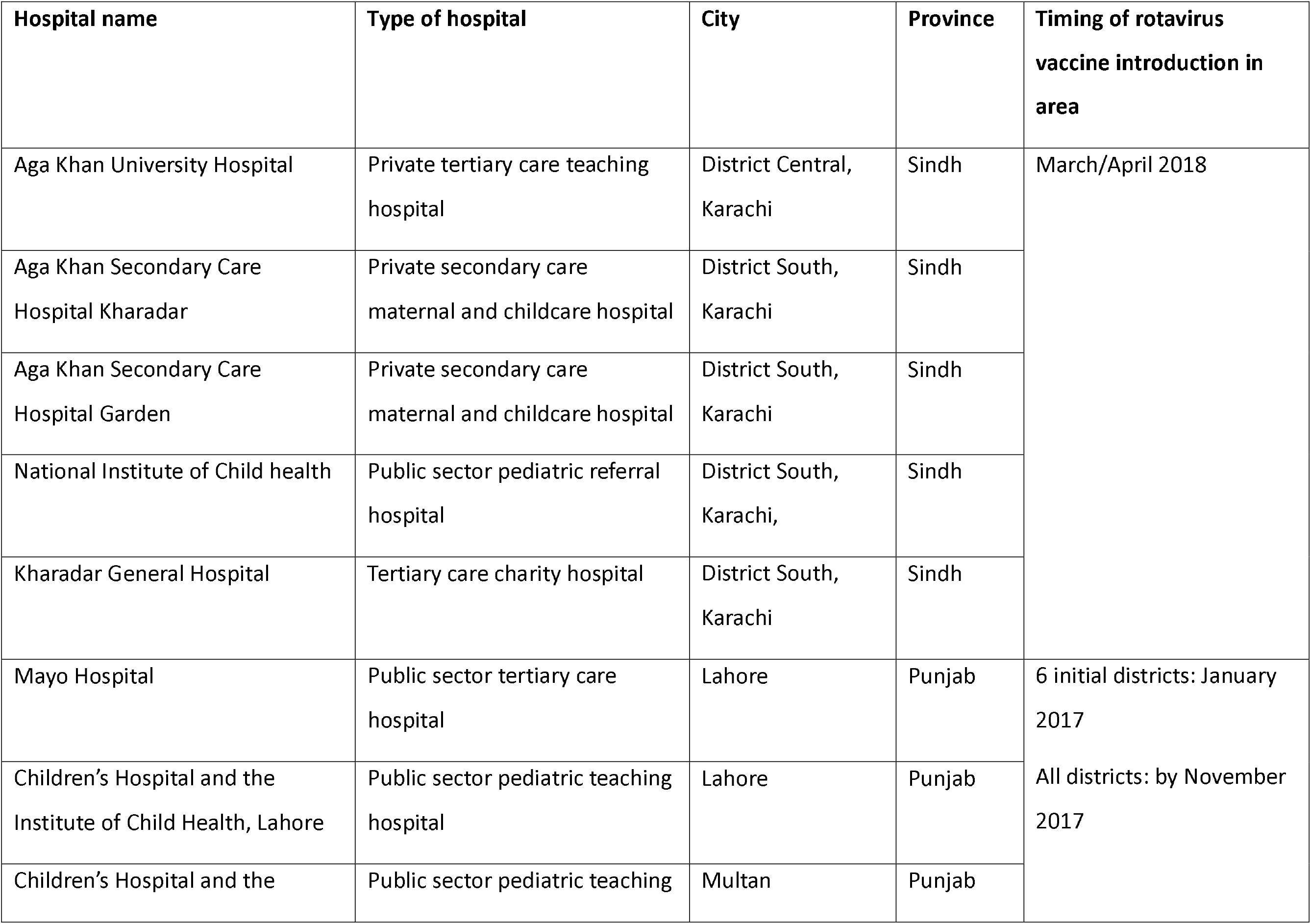
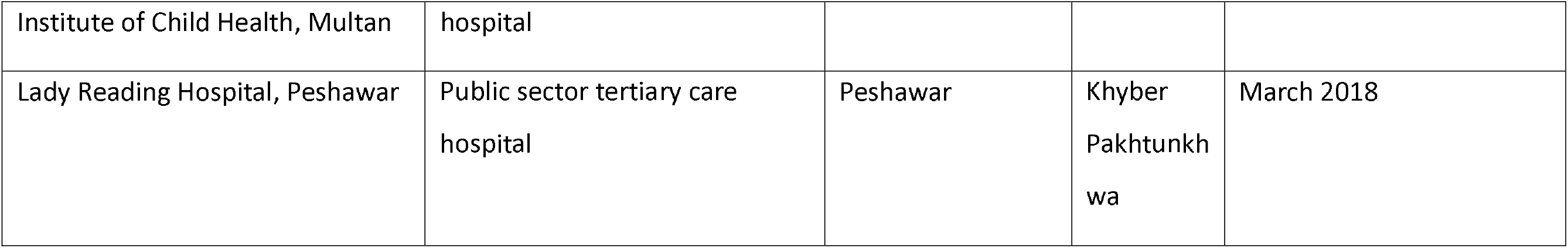
Location and information on participating hospitals - Pakistan, 4/2018 – 3/2023

**Figure 2.**
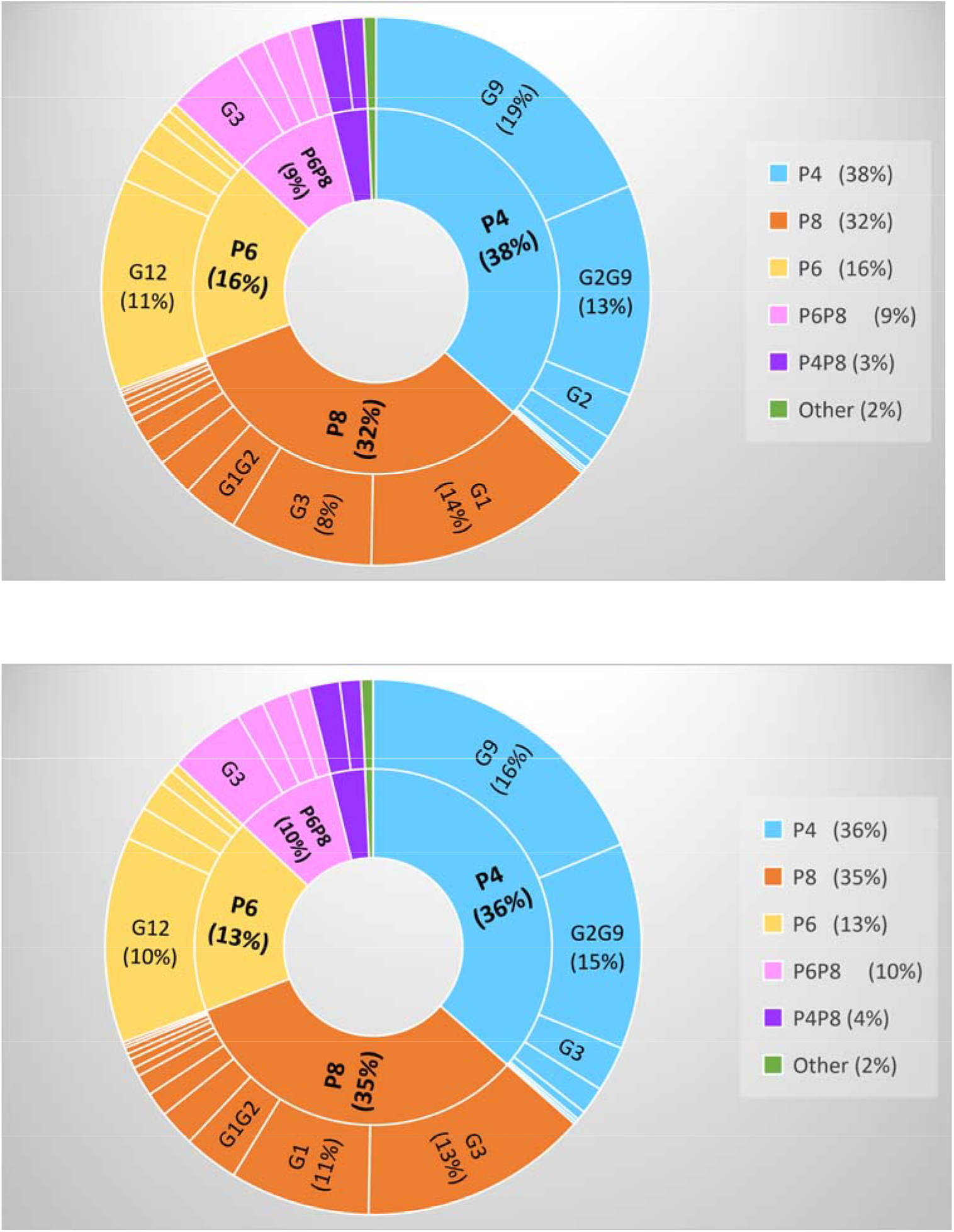
Distribution of rotavirus genotypes among rotavirus cases used in the vaccine effectiveness analysis of 2 doses of RV1 vs 0 doses, by age group - Pakistan, 4/2018 – 3/2023 (top figure: children aged 22 weeks−11 months, n=485; bottom figure: children aged 12 months−23 months, n=209). Excludes cases from one facility (see text).

Excluding the 26 cases and 129 controls from Mayo Hospital aged 22 weeks−11 months, the overall 2 dose VE of 33% (95% CI 12, 50) was nearly identical to the overall VE for this age group (Table 1). Because case counts in categories by separate G- and P-type combinations, or by single G-types, were small for most categories, we examined VE by P genotype among cases where only 1 P genotype was detected. Examining 2 dose VE for those aged 22 weeks−11months, VE against disease with only P[8] detected was 36% (CI 0, 59); with only P[4] detected was 21% (95% CI -21, 48) and with only P[6] was 56% (95% CI 25, 74) (Table 4). The 2 dose VE against disease with P[4] NOT detected (i.e., those with only P[8] and/or P[6] detected, or P-nontypeable) was 43% (95% CI 20, 59). The VE against disease with only G1P[8] detected was 19% (95% CI -53, 57), based on a small number of cases (n=66); 31 (47%) of these had been tested for RV1 strain and none were positive. Among the individual P-type cases, distribution of severity scores did not explain potential differences in point-estimates (data not shown). When genotype analyses among those aged 22 weeks−11 months were stratified by WAZ category, the VE point estimates were higher among those with score ≥-2, compared to those with score <-2, but the 95% CI often included zero (Table 4).

**Table 4.**
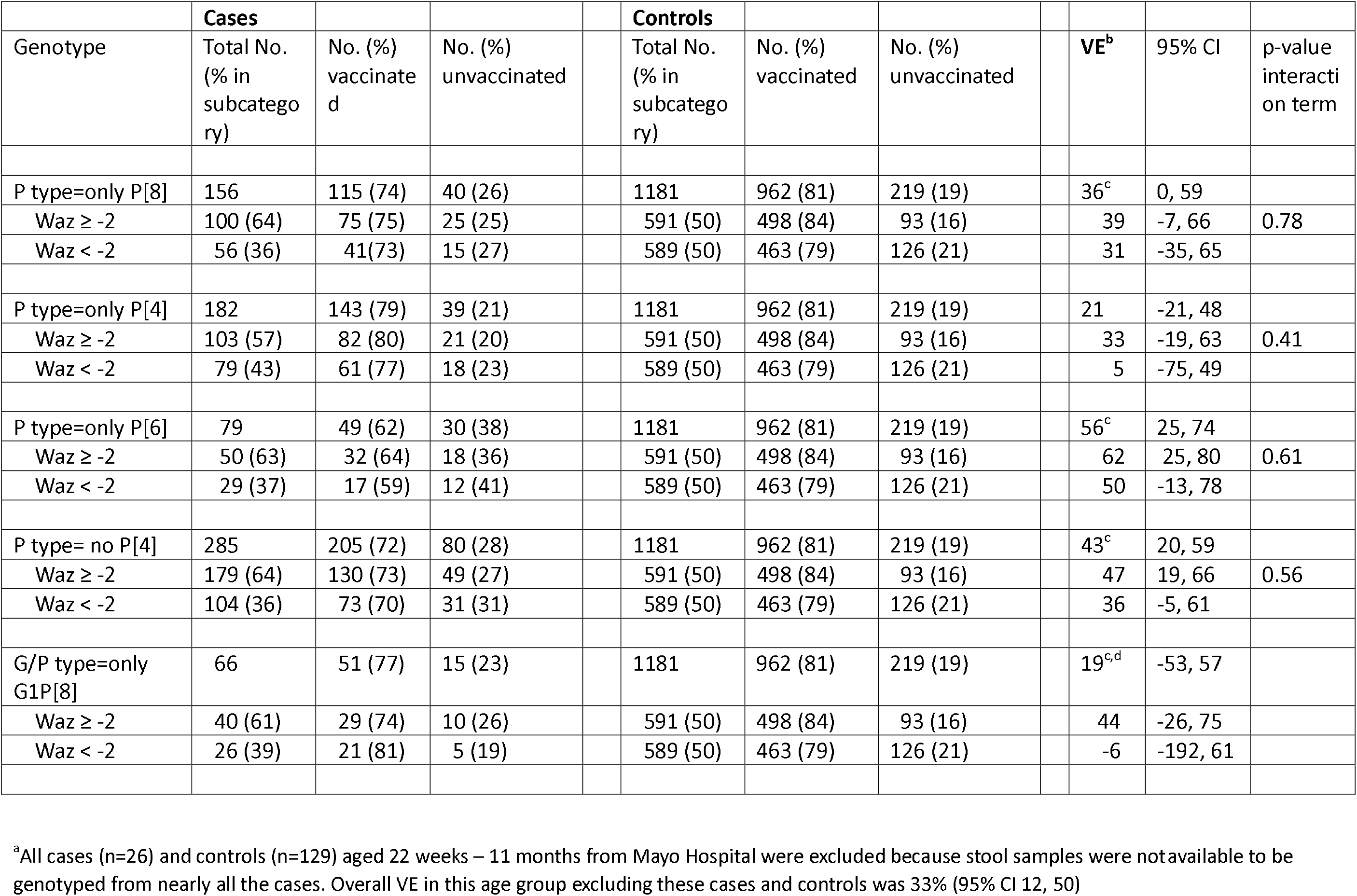
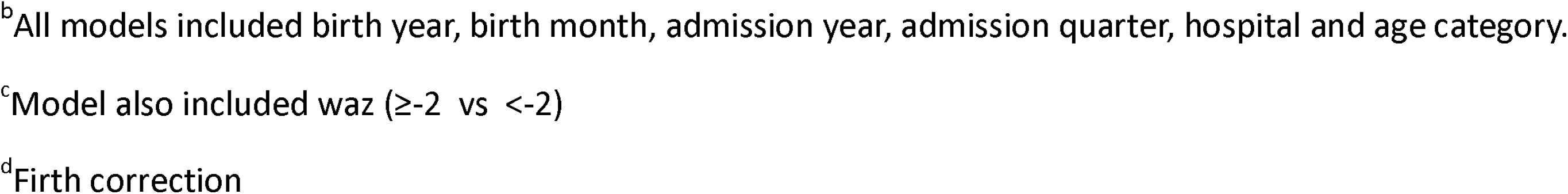
Vaccine effectiveness of 2 doses of RV1 vs 0 doses in children aged 22 weeks−11 months, by genotype^a^ - Pakistan, 4/2018 – 3/2023.

|  | Cases |  |  |  | Controls |  |  |  |  |  |  |
| --- | --- | --- | --- | --- | --- | --- | --- | --- | --- | --- | --- |
| Genotype | Total No.<br>(% in<br>subcategory) | No. (%)<br>vaccinated | No. (%)<br>unvaccinated |  | Total No.<br>(% in<br>subcategory) | No. (%)<br>vaccinated | No. (%)<br>unvaccinated |  | VE<sup>b</sup> | 95% CI | p-value<br>interaction term |
| P type=only P[8] | 156 | 115 (74) | 40 (26) |  | 1181 | 962 (81) | 219 (19) |  | 36<sup>c</sup> | 0, 59 |  |
| Waz ≥ -2 | 100 (64) | 75 (75) | 25 (25) |  | 591 (50) | 498 (84) | 93 (16) |  | 39 | -7, 66 | 0.78 |
| Waz < -2 | 56 (36) | 41(73) | 15 (27) |  | 589 (50) | 463 (79) | 126 (21) |  | 31 | -35, 65 |  |
| P type=only P[4] | 182 | 143 (79) | 39 (21) |  | 1181 | 962 (81) | 219 (19) |  | 21 | -21, 48 |  |
| Waz ≥ -2 | 103 (57) | 82 (80) | 21 (20) |  | 591 (50) | 498 (84) | 93 (16) |  | 33 | -19, 63 | 0.41 |
| Waz < -2 | 79 (43) | 61 (77) | 18 (23) |  | 589 (50) | 463 (79) | 126 (21) |  | 5 | -75, 49 |  |
| P type=only P[6] | 79 | 49 (62) | 30 (38) |  | 1181 | 962 (81) | 219 (19) |  | 56<sup>c</sup> | 25, 74 |  |
| Waz ≥ -2 | 50 (63) | 32 (64) | 18 (36) |  | 591 (50) | 498 (84) | 93 (16) |  | 62 | 25, 80 | 0.61 |
| Waz < -2 | 29 (37) | 17 (59) | 12 (41) |  | 589 (50) | 463 (79) | 126 (21) |  | 50 | -13, 78 |  |
| P type= no P[4] | 285 | 205 (72) | 80 (28) |  | 1181 | 962 (81) | 219 (19) |  | 43<sup>c</sup> | 20, 59 |  |
| Waz ≥ -2 | 179 (64) | 130 (73) | 49 (27) |  | 591 (50) | 498 (84) | 93 (16) |  | 47 | 19, 66 | 0.56 |
| Waz < -2 | 104 (36) | 73 (70) | 31 (31) |  | 589 (50) | 463 (79) | 126 (21) |  | 36 | -5, 61 |  |
| G/P type=only G1P[8] | 66 | 51 (77) | 15 (23) |  | 1181 | 962 (81) | 219 (19) |  | 19<sup>c,d</sup> | -53, 57 |  |
| Waz ≥ -2 | 40 (61) | 29 (74) | 10 (26) |  | 591 (50) | 498 (84) | 93 (16) |  | 44 | -26, 75 |  |
| Waz < -2 | 26 (39) | 21 (81) | 5 (19) |  | 589 (50) | 463 (79) | 126 (21) |  | -6 | -192, 61 |  |

<sup>a</sup>All cases (n=26) and controls (n=129) aged 22 weeks – 11 months from Mayo Hospital were excluded because stool samples were not available to be genotyped from nearly all the cases. Overall VE in this age group excluding these cases and controls was 33% (95% CI 12, 50)
<sup>b</sup>All models included birth year, birth month, admission year, admission quarter, hospital and age category.
<sup>c</sup>Model also included waz ( $\geq -2$ vs $< -2$ )
<sup>d</sup>Firth correction

Among cases aged 12−23 months used in the 2 dose VE analysis (excluding cases and controls from Mayo Hospital), the distribution of genotypes was similar to that of children aged 22 weeks−11 months (Figure 2, Supplemental Table 6) and similarly, 39% (80/207 with at least G- or P-typeable) of cases had >1 G and/or P type detected. For this age group, separate P-genotype categories generally had low case numbers, and VE calculations had low point estimates and wide CIs that included zero (Supplemental Table 7).

## DISCUSSION

In this test-negative case-control evaluation of effectiveness of 2 doses of oral RV1 vaccine against rotavirus disease with hospital-based care involving data from over 2900 systematically enrolled children at 9 facilities, we estimated VE among infants aged 22 weeks–11 months to be 33% (95% CI 12, 49) with higher point estimate of 45% (95% CI 21, 62) among those who were better nourished. Among children aged 1 year, VE estimate was 24% (95% CI -20, 52), with confidence interval that included zero. There was a wide array of genotypes detected amongst our cases and nearly 40% of cases had >1 G and/or P genotype identified.

Strengths of our evaluation include large numbers of children enrolled, anthropomorphic measurements obtained near discharge and genotyping results from nearly all rotavirus cases. Substantial effort was made to continue enrollment during most of the COVID-19 pandemic period and appeared generally successful. These efforts allowed for stratified analysis by subgroups of interest to aid in understanding factors that may impact VE results.

Overall, our point estimate for VE in age <1 year is lower than other VE point estimates for RV1 in other countries with high under 5 mortality rates; a random-effects model estimates VE for age <12 months as 63 (95% CI 54, 70) from 10 evaluations published 2013−2019 [14]. Our point estimate was within the 95% CI (near the lower limit) for 8 of the individual evaluations. VE evaluations in this age group from Central Asian countries with high under-5 mortality rates published subsequently include from one hospital in Uzbekistan (VE 52% [95% CI -40, 84] and one in Tajikistan (VE 61% [95%CI 17-81] [15,16]. Our point estimate for infants is also lower than that from our likely most comparable setting, Afghanistan, conducted at 4 hospitals during 2018−2021 (VE for age 6−11 months: 57% [95% CI 33, 73]) which was based on a similar number of cases and controls as ours [17]. In Afghanistan, the VE for this age group against illness with modified Vesikari score ≥11 was 46% (95% CI 11, 67), which is similar to point estimates found in RV1 trails in Bangladesh and Malawi [18, 19]. Comparison of results from observational evaluations across high-mortality countries (or even within one country) requires some caution given expected differences in enrollment settings, limitations of severity measurements, ability to accurately capture vaccination status, health care seeking behavior, nutritional status, unmeasured confounders or effect modifiers (including histo-blood group antigens [HBGA] of the infants and mothers, and perhaps very importantly, background rate of infection [20] and probably circulating genotypes.

Among children aged 12−23 months, our VE estimate was low and statistically non-significant; our VE for age 22 weeks−23 months, however, retained statistical significance. In the random effects analysis described above, the VE in the 12−23 month age group was 58% (95% CI 38, 72); the 95% Cis included zero for 3 of the 5 individual evaluations [14]. Our results for age 12−23 months are similar to those from Afghanistan (18 [95% CI -49, 59] and the Bangladesh and Malawi trials [17-19]. Lower vaccine efficacy or effectiveness in the second year of life has been frequently reported with rotavirus vaccines in high under-5 mortality countries. While further modeling efforts have confirmed this not unexpected finding, they have importantly identified that our usual measurement approach can overestimate the degree of waning in settings with high forces of infection [21]. Our overall crude versus adjusted VE estimate indicated confounding was present in this age group; residual confounding may still be present and estimates in this age group may be particularly sensitive to variables included. Different from what some assessments found among children in low income countries, we did not observe a general trend toward higher VE against disease with higher modified severity score (which relies in large part on parental report of symptoms), but point estimates appeared higher in a few categories.

According to the 2018 Pakistan National Nutritional Survey, 26% of infants aged 6−11 months were underweight (WA z-score<-2) and 32% were stunted (HA z-score<-2); these proportions were higher in children aged 1 year (underweight: 25%-30%; stunted 39%-47%) [13]. Compared to the 2018 survey results, higher proportions of children in our evaluation (who presented for acute gastroenteritis) were underweight or stunted. Among infants, our overall VE point estimate (and VE point estimate for some genotypes) was higher among infants who were better nourished compared to those malnourished as assessed by weight-for-age. On the other hand, our VE estimates were very similar among stunted and non-stunted infants (modest difference in point estimate when using separate models). It is possible that, in our evaluation, weight was generally a more accurate measurement than was length. It is likely that that nutritional status at the time of vaccine receipt as well as at the time of exposure to wild-type rotavirus both contribute to the outcome from rotavirus exposure. Among our children aged 1 year, confidence intervals included zero in all nutritional categories. As summarized by Burnett et al, RV1 VE estimates in high mortality countries that also examined VE by nutritional status had overall low case numbers in the malnourished groups (except for the Zimbabwe evaluation) and usually included non-infants [22-26]. While the nutritional measurements used varied, those results overall supported lower VE point estimates in malnourished children [17,22].

Because RV1 contains only an attenuated G1P[8] strain, there has long been questions about whether it protects well against disease from genotypes that share neither G nor P type with RV1 (“fully heterotypic”), particularly G2P[4, and especially in high mortality countries [27-8]. G2P[4] had been a prevalent strain world-wide with all 11 genes (based on a DS-1 genetic “backbone”) different from that of RV1 strain (Wa-like backbone). Pooling data from all RV1 efficacy trials for robust analyses, Amin et al identified that efficacy of RV1 was indeed lower against G2P[4] disease compared with that against G1P[8]; this lower efficacy for fully heterotypic strains was maintained when P[6] strains (about a quarter of the fully heterotypic strains) and other non-G1 non-P8 strains were included [29]. Most of our cases had genotype(s) fully heterotypic from RV1. In our infant group, our VE point estimate against disease with only P[4} detected appeared lowest, not only compared with the only-P[8] disease estimate but also compared with the only-P[6] estimate (our highest point estimate). Malawi investigators found RV1 VE estimates were similarly high against P[8] and P[6] strains, and lower against P[4] disease with confidence interval that included zero; in their evaluation, most P[4] strains were G2P[4] [26]. Surprisingly, our VE against disease with only G1P[8] detected was low overall, although this was based on a smaller number of cases. VE estimates using classifications based on additional molecular analyses of strains (rather than only overall G,P type) may provide further insights. For example, G1P[8] strains with a DS-1 backbone have been identified in the past decade; with a small number of cases, investigators in Malawi assessed that RV1 maintained effectiveness against this strain [30]. Interpretation of overall and genotype-specific vaccine efficacy/effectiveness results (and comparison across evaluations) may be further complicated by the populations’ HBGA of infants and mothers, which are believed to impact rotavirus susceptibility in a P-type dependent manner [31-5].

Our evaluation has several limitations. As in all similar evaluations without availability of an accurate electronic vaccination registry, confirming that a particular child indeed had not received any doses of rotavirus vaccine is challenging. We relied largely on verbal report from mother that the child had not been vaccinated with any vaccines subsequent to any birth doses. Overall, we were not able to obtain rotavirus vaccination information in 17% of infants and 24% of 1 year olds. While our project was designed to have weight measured at discharge to better represent the child’s pre-morbid weight, it is likely that some weight measurements did not accurately reflect this status. We enrolled large numbers of children so that we would have sufficient cases to estimate VE stratified by certain characteristics, but we were not specifically powered for each outcome nor to identify statistically significant differences between strata, if they indeed existed. Although planned a priori, we performed a large number of subanalyses, increasing the possibility of spurious findings, and some categories had relatively small case numbers which may result in inaccuracies. Further, residual unmeasured confounding from a high background rotavirus infection rate may have biased VE estimates low, including in our infants. (*Amin2*). Lastly, the impact of the COVID-19 pandemic on our results or on our ability to reliably estimate effectiveness is unknown.

In our settings in Pakistan, which are likely among those with the greatest hurdles for an oral vaccine given in early infancy, our evaluation supports that 2 doses of RV1 given in the routine immunization program along with OPV provides fair protection against rotavirus disease resulting in hospital care; with high vaccine coverage, the overall impact of vaccination on rotavirus morbidity and mortality can be important. Our findings support the value of continued efforts to improve rotavirus vaccine performance in such settings, which include assessing an additional dose in later infancy (underway in Pakistan), ongoing effectiveness evaluations of unique products (i.e., those using neonatal strains [Rotavac and RVBB3] and neonatal doses [RVBB3]), and ultimately the addition of an injectable vaccine to oral vaccine regimens [36-9].

## Data Availability

All data produced in the present study may be made available upon reasonable request to the authors

## Disclaimer

The findings and conclusions in this report are those of the authors and do not necessarily represent the official position of the US Centers for Disease Control and Prevention.

## Author contributions

### Conceptualization and Methodology

Syed Asad Ali, Umesh Parashar, Jacqueline Tate, Margaret Cortese

### Investigation

Shazia Sultana, Atif Riaz, Aneeta Hotwani, Furqan Kabir, Muhammad Nasir Rana, Yasin Alvi, Saadia Khan, Khalid Iqbal, Mubashir Ahmed, Muhammad Haroon Hamid, Amir Muhammad, Inayat Ullah Afridi, Arit Parkash, Nasir Saleem Saddal, Jamal Raza, Muhammad Hayat Bozdar, Zafar Iqbal Channa, Zareef Uddin Khan, Muhammad Naeem Rajput, Sohail Raza Shaikh

### Formal Analysis

Margaret Cortese

### Data Curation

Shazia Sultana, Atif Riaz, Margaret Cortese

### Writing: original draft

Mohammad Tahir Yousafzai, Margaret Cortese

### Writing: review and editing

All co-authors

### Supervision

Syed Asad Ali, Shazia Sultana, Atif Riaz, Mathew Esona, Rashi Gautam, Umesh Parashar, Muhammad Ahmed Kazi

### Project administration

Shazia Sultana, Atif Riaz, Muhammad Ahmed Kazi Funding acquisition: Syed Asad Ali

---

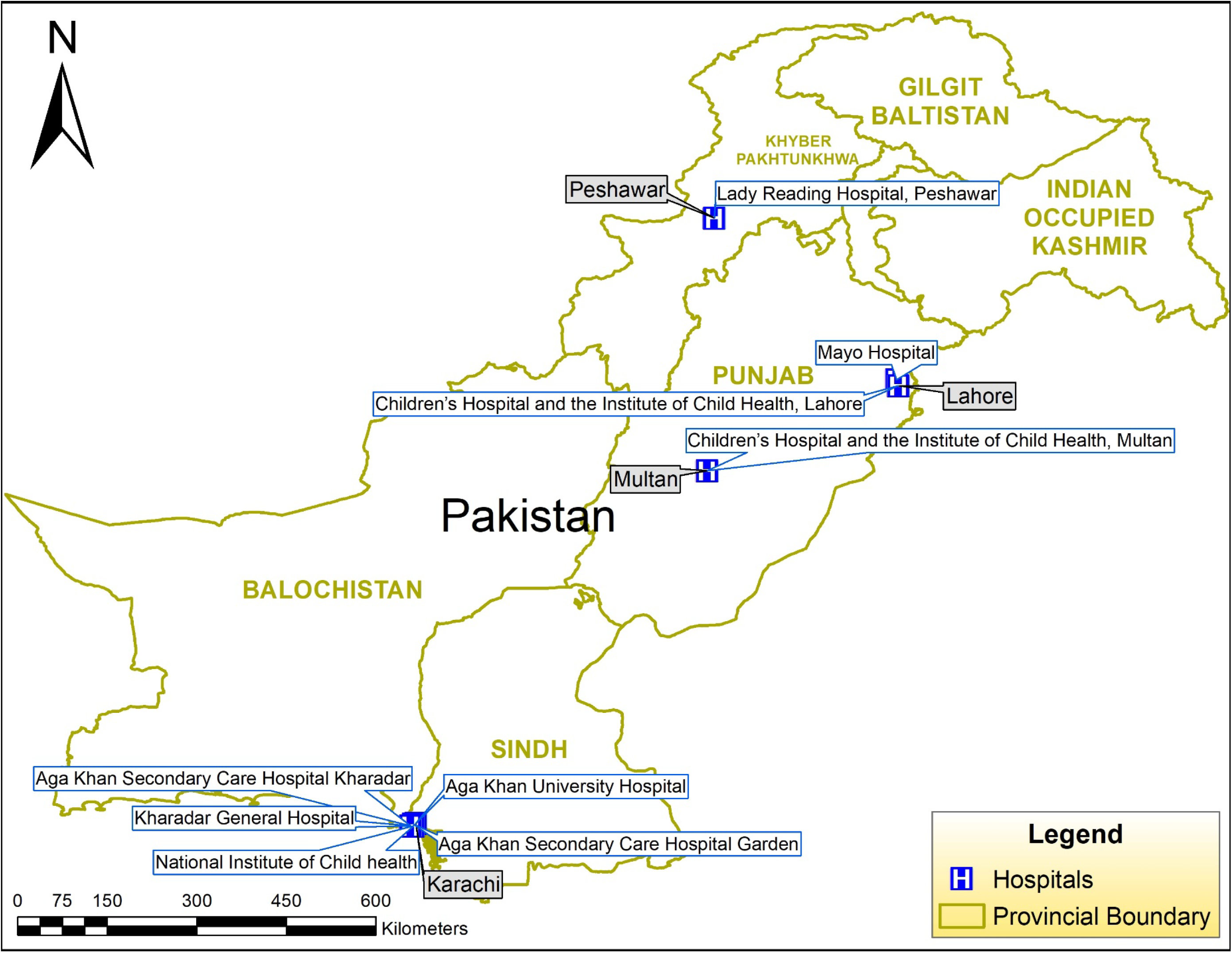

